# Health coverage for people without social security in Mexico: a retrospective cohort to assess childhood acute lymphoblastic leukaemia survival

**DOI:** 10.1101/2020.07.09.20149302

**Authors:** P Muñoz-Aguirre, R Huerta-Gutierrez, S Zamora, A Mohar, L Vega-Vega, JE Hernández-Ávila, E Morales-Carmona, M Zapata-Tarres, S Bautista-Arredondo, R Perez-Cuevas, R Rivera-Luna, MR Reich, M Lajous

## Abstract

We aimed to measure survival of children with acute lymphoblastic leukaemia (ALL) under Mexico’s public health insurance for the population without social security, and explore patient groups that may be at risk for increased mortality.

We conducted a retrospective cohort study using claims data from Mexico’s *Seguro Popular* program, covering cancer treatment in people without social security, 2005-15. We estimated 5-year national and state-specific survival for children with ALL across Mexico who initiated cancer treatment under this public healthcare insurance scheme.

8,977 children with ALL were treated under *Seguro Popular* in 2005-15. Under this financing scheme, treated children doubled from 535 in 2005 to 1,070 in 2015, and their 5-year survival was 61.8% (95%CI 60.8, 62.9). Estimates for 5-year survival remained constant over time. We observed wide gaps in risk-standardized 5-year survival among states ranging from 74.7% to 43.7%. We found a higher risk of mortality for children who received treatment in a non-paediatric specialty hospital (Hazards Ratio, HR=1.18; 95%CI 1.09, 1.26), facilities without a paediatric oncology/haematology specialist (HR=2.17; 95%CI 1.62, 2.90), and hospitals with low patient volume (HR=1.22; 95%CI 1.13, 1.32).

In a decade Mexico’s *Seguro Popular* doubled access to ALL treatment for children without social security and by 2015 financed the vast majority of estimated ALL cases for that population. While some progress in ALL survival may have been achieved, nationwide 5-year survival was unsatisfactory and did not improve over time.

Our results provide lessons for Mexico’s evolving health system and for countries moving towards universal health coverage.

**Novelty and impact:** There has been no comprehensive assessment of the health outcomes of Mexico’s policy for financing high-cost cancer treatments that was part of *Seguro Popular*, health reform aimed at providing care to the population without social security. The study identified several factors amenable to intervention that could improve survival for childhood ALL in Mexico and reduce the observed disparities.

Decision could consider administrative databases as a source of clinical outcomes to ensure the quality their systems.

## Introduction

As countries move towards universal health coverage, improving our understanding of financing in health and effectiveness of healthcare delivery systems has become a global priority.^1^ *Seguro Popular*, Mexico’s 2003 landmark health reform, financed health coverage to the population without social security (roughly, 57% of Mexico’s population) between 2004 and 2019.^2^ While childhood cancer represents a small proportion of cancers, it is an important priority for many countries. According to CONCORD-3, most developed countries have achieved significant progress in the diagnosis, treatment, and supportive care for the most common childhood cancer, acute lymphoblastic leukaemia (ALL), with 5-year survival rates approaching 90%.^3^ In Mexico, *Seguro Popular* recognized childhood cancer as a priority and provided access to care for all children with cancer in the population without social security by financing oncology treatments under specific guidelines.^4^

In Latin America, Argentina and Chile have demonstrated significant advances in cancer care by achieving a 70% 5-year survival of children with ALL.^5 6^ In Mexico, while no reliable national estimates prior to *Seguro Popular* are available, an initial assessment of this policy found increasing coverage of childhood cancer care with an estimated survival of 50% for ALL.^7^ As Mexico’s current government restructures the health system^8 9^ and integration of non-communicable diseases to universal health coverage frameworks are considered for other countries,^10^ a comprehensive assessment of the health outcomes achieved under Mexico’s financing policy for high-cost cancer treatments is warranted.

We evaluated the effectiveness of *Seguro Popular’s* financing policy by exploring childhood ALL survival between 2005 and 2017 in Mexico and across states, and examined patient groups that may be at risk for increased mortality. We use Mexico’s experience to draw lessons for improving cancer care delivery.

## Methods

### Study design

We conducted a retrospective cohort study using nationwide comprehensive reimbursement data from the Fund for Protection against Catastrophic Expenses (FPGC, for its Spanish acronym) as part of *Seguro Popular*, which was managed by Mexico’s National Commission for Social Protection in Health (CNPSS, for its Spanish acronym). Until December 2019, CNPSS was the agency responsible for the implementation and operation of *Seguro Popular* which covered through the FPGC a specified list of 65 high-cost interventions, including (standard and high risk) childhood ALL. These patients received risk-based treatment protocols defined by an expert group of paediatric oncologists and validated by national health authorities. We included in our analysis patients registered for reimbursement for this disease between January 2005 and December 2015. We excluded patients in relapse and those without a plausible date for treatment initiation or older than 18 years of age (**Supplementary Figure 1**). The project was approved by the National Institute of Public Health’s Institutional Review Board (P121-18/CI:1586). This study was funded in part by Fundación Gonzalo Rio Arronte and Mexico’s Consejo Nacional de Ciencia y Tecnología. The study sponsors had no role in the data collection, analysis, or interpretation of results.

## Data sources

### Reimbursement data

Details on FPCG reimbursement data management and harmonization are included in the **Supplementary Appendix 1**. Briefly, we explored yearly datasets, consulted with CNPSS personnel and clinicians to harmonize variables, and created a single analytic database. Before January 1, 2011, there was one reimbursement claim per patient. Beginning on that date the reimbursement method changed to fee-for-service. Thus, multiple claims (observations) per individual were possible. We identified 29,005 reimbursement claims for the study period representing 9,535 children (**Supplementary Figure 1**). After exclusions, we classified patients according to the year of treatment, risk (standard/high), state of residence and treatment, whether they migrated for treatment, and the type of hospital (general or paediatric specialty/with or without a paediatric haematologist or oncologist/low, medium, or high patient volume according to tertiles of children treated in the previous year).

We assessed the completeness and quality of reimbursement data comparing children in our study to those included in a nationwide retrospective medical record review of ALL patients without social security in Mexico treated between January 2006 and September 2009.^7^ We found 81.8% children from that review in our reimbursement dataset. Unfortunately, our capacity to complete cross-linkage was limited because a valid national identification number was not available for the remaining children. Also, for the most part the information recorded in the reimbursement claims corresponded to what was reported in medical records (**Supplementary Appendix 2**).

### Mortality data linkage

We cross-linked study participants to Mexico’s Epidemiological and Statistical System of Mortality (SEED), a mortality registry designed for disease surveillance and maintained by the Ministry of Health. Using a previously validated record linkage algorithm, we identified deaths occurring between January 2005 and December 2017 (the last available date when mortality data was available**; Supplementary Appendix 1**).^11^ We assessed the validity of our mortality linkage strategy by searching 20 children who died before December 31, 2017, and 40 children known to be alive after that date who received cancer treatment at one of the accredited providers (*Hospital Infantil Teletón de Oncología* or HITO). All deaths were identified in the mortality registry and none of the children known to be alive after 2017 were found (i.e., perfect sensitivity and specificity). Among the 20 deaths the cause of death was solid tumour for nine children, central nervous system tumours for five, ALL for four, and two deaths were attributable to cancer treatment complications.

## Statistical analyses

First, to describe how the financing policy was implemented we determined the distribution of ALL patients according to year of treatment, state of residence, and state of the treating hospital. Because remoteness to paediatric oncology facilities affects clinical outcomes,^12 13^ we estimated the overall and state-specific proportion of patients who travelled out-of-state to seek treatment.

Second, to estimate ALL survival, patients were followed-up from the date when treatment began until death or December 31, 2017. We used the Kaplan-Meier method to compute the survival function and estimated total 1-, 2-, 3-, 4-, and 5-year percent survival. We also estimated 30-day survival (or induction mortality) because patients may experience toxicity and treatment-related mortality in the induction phase and reducing early mortality in ALL is an important goal to improve overall survival.^14^ We evaluated changes in survival over time by estimating 5- and 3-year percent survival according to year of reimbursement (2005-2014). We calculated 95% confidence intervals (95% CI) using log-log transformation and used the Breslow method to handle ties. For comparability between states and providers we standardized survival estimates by risk according to the national distribution of risk categories.

Finally, to identify patient groups at risk for increased mortality, we compared survival curves of patients who sought treatment outside of their state of residence (migrated) to those who remained in their home states. We also evaluated the relative survival of patients treated in a general specialty hospital (vs. paediatric specialty hospital), a hospital without a paediatric oncology/haematology specialist (relative to a facility with such a specialist), and a low or medium patient-volume hospital (vs. a high patient-volume). Comparisons among groups were conducted using the log-rank test with a significance level of 0.05. We estimated multivariable-adjusted mortality hazard ratios and 95% CIs based on subject-matter knowledge using Cox regression. We assumed proportionality of the hazards and censoring at random and verified the proportional hazards assumption plotting the Schoenfeld residuals against time. Data management, analyses, and data visualization were conducted using SAS software (Version 9.4, SAS Institute Inc.) and R (R Core Team, 2019).

## Results

Between January 1, 2005, and December 31, 2015, we identified 8,977 children with ALL, whose treatment was approved for reimbursement through FPGC. The number of treated patients rose steadily from 535 in 2005 to 1,070 in 2015 (**Figure 1**). Over the study period six states treated close to half (48.4%; **Supplementary Table 1**) of ALL patients and the proportion of patients who migrated to receive treatment (i.e., treated outside their state of origin) progressively declined from 20.9% (112 out of 535 patients treated that year) in 2005 to 16.4% (or 175 out of 1,070 patients) by 2015.

**Figure 1.**
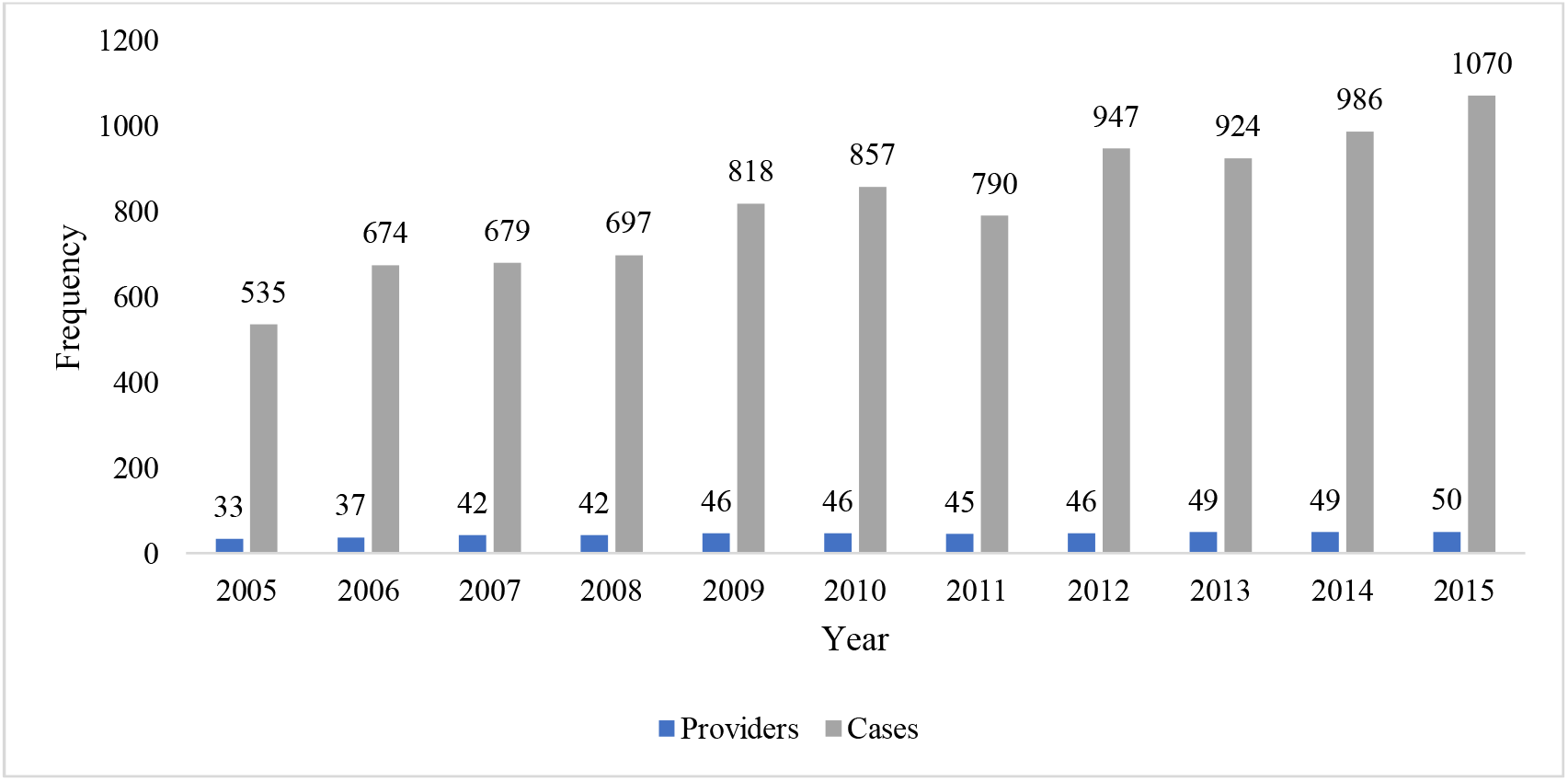
Certified hospitals for treatment of childhood ALL and treated cases per year under *Seguro Popular*’s policy for financing high-cost treatments, 2005-2015.

The mean (±SD) age of patients was 7.4 (4.7; **Table 1**) and the proportion of patients with high-risk ALL was 69.5%. The median patient volume (number of patients treated in the previous year) was 26 (ranging from 1 to 82). Most children were treated in their state of residence (83.5%) and in a non-paediatric general specialty hospital (55.6%). The mean days from diagnosis to treatment was 7 days with a range of 3 to18. The range in the number of ALL cases treated in the previous year in hospitals considered to be of high patient volume was 36 to 82. Only 63 patients were treated in a facility without a paediatric specialist. Overall, patient characteristics did not appear to change importantly over the years (**Supplementary Table 2**). However, the proportion of individuals with high-risk ALL appeared to increase over time.

**Table 1.** Characteristics of children with ALL treated under *Seguro Popular*’s policy for financing high-cost treatments, 2005-2015.

| **Patient characteristics\*** | **n = 8,977** |
| --- | --- |
| Girls | 4,088 (45.5) |
| Age at diagnosis in years, mean (±SD) | 7.4 (4.7) |
| Age groups in years |  |
| 0-5 | 4,005 (44.6) |
| 6-10 | 2,386 (26.6) |
| 11-15 | 2,019 (22.5) |
| 16-18 | 597 (6.3) |
| High risk | 6,236 (69.5) |
| Days from diagnosis to treatment (median, interquartile range)** | 7 (3-18) |
| Treatment |  |
| Outside state of residence | 1,481 (16.5) |
| Non-paediatric specialty hospital | 4,992 (55.6) |
| Hospital without a paediatric specialist | 63 (0.7) |
| Hospital patient volume, patients per year |  |
| Low, 1-19 | 3,057 (34.0) |
| Medium, 20-35 | 2,889 (32.2) |
| High, 36-82 | 3,031 (33.8) |

\*Frequency (%), unless specified; \*\*Missing values for 1,040 (11.6%)

Median follow-up was 3.9 years (interquartile range: 1.9, 7.5) and 42,751 person-years were accrued over the study period. Five-year survival for childhood ALL cases reimbursed by *Seguro Popular* between 2005 and 2015 was 61.8% (95%CI 60.8, 62.9; **Figure 3**) and 64.0% (95%CI 62.9, 65.1) for children 0-14 years. Overall 5.3% of children died within 30 days (early mortality), which represents the induction phase of treatment (30-day survival of 94.7%; 95%CI 94.2, 95.1). Estimates for 5-year survival remained constant over time. For 2005, 5-year survival was 60.9% (95%CI 56.7, 64.9) and for 2012, it was 61.4% (95%CI 58.2.1, 64.4). Similarly, when we repeated analyses for 3-year survival we did not observe changes over time (**Supplementary Table 3**). In 2011, the reimbursement method changed from bundled (per case) payment to fee-for-service payment. Thus, we compared survival before and after that year and found no difference (61.5%; 95%CI 60.0, 62.9 before 2011 vs. 62.1%; 95%CI 60.6, 63.6 after 2011). Five-year survival was 72.5% (95%CI 70.3, 74.2) for standard-risk and 56.9% (95%CI 55.6, 58.2) for high-risk patients. Estimates were similar after exclusion of children for whom no justification was provided by the treating hospital **(Supplementary Table 4)**.

**Figure 2.**
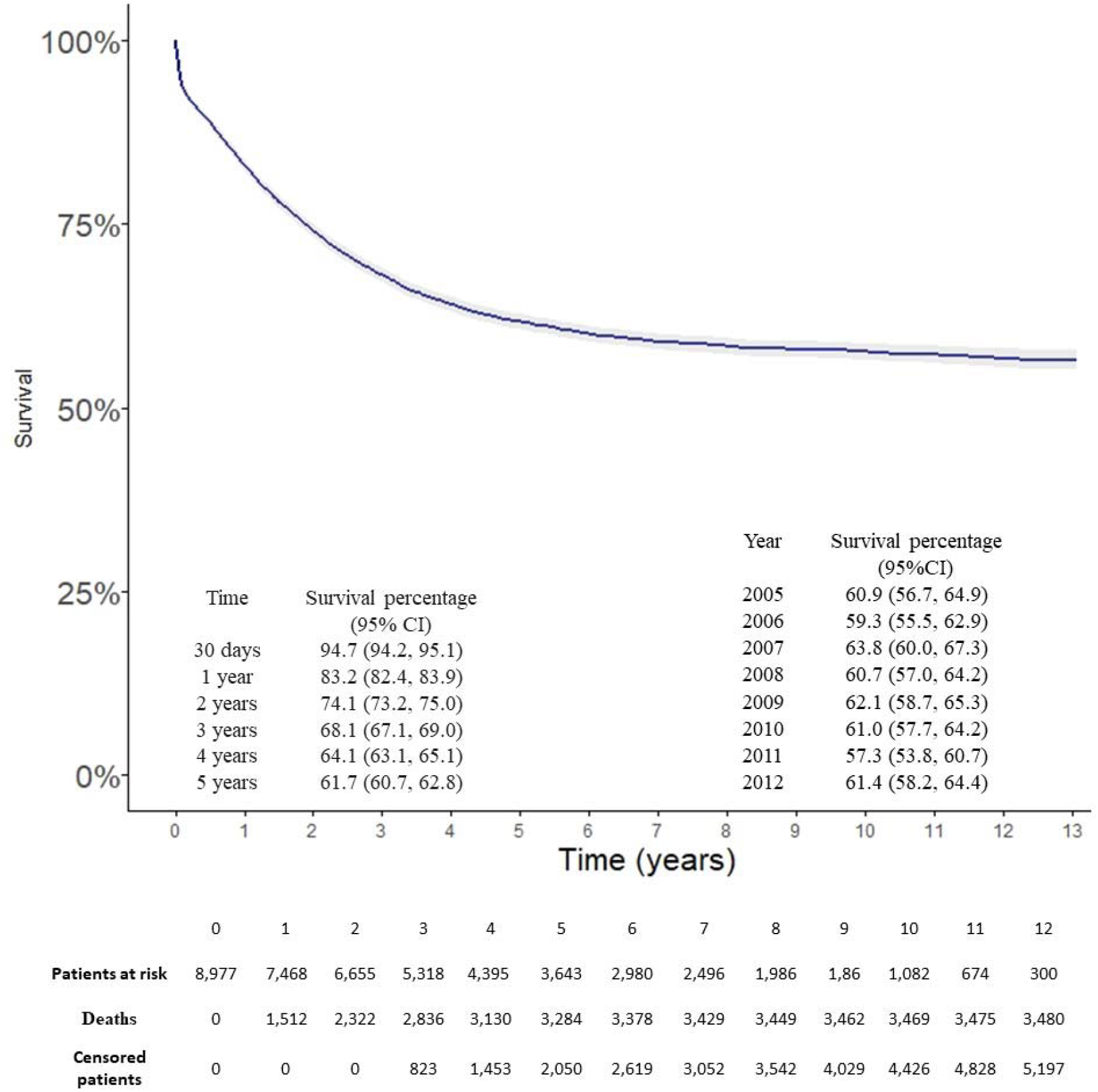
Overall 5-year survival among 8,977 children with ALL treated under *Seguro Popular*’s policy for financing high-cost treatments, 2005-2015.

**Figure 3.**
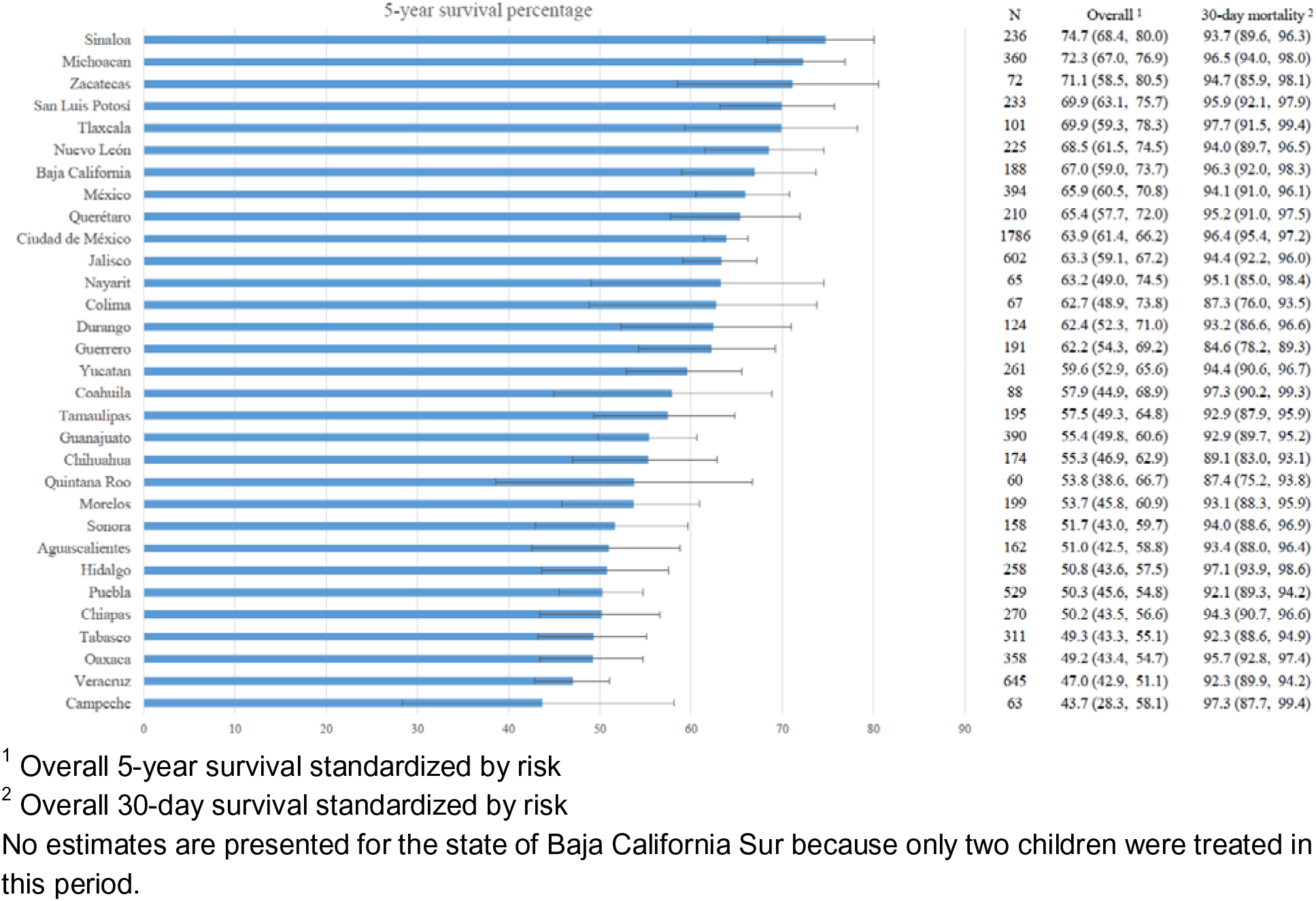
Overall 5-year and early survival, and 95% confidence intervals for children with ALL treated under *Seguro Popular*’s policy for financing high-cost treatments by state, 2005-2015.

We observed important heterogeneity in crude survival across states (**Supplementary Table 5**), which remained even after accounting for differences in the distribution of risk categories through standardization (**Figure 4**). While 5-year risk-standardized survival was above 70% for Sinaloa, Michoacán, and Zacatecas, there were six states where survival was 50% or below. We found that early mortality, possibly representing oncologic emergencies, toxicity, and treatment-related mortality in the induction phase (induction mortality), also differed importantly across states. There were only 7 states where induction mortality was less than 4%. These disparities remained when we explored survival according to state of origin (**Supplementary Figure 2**) or limited the analysis to children treated in their states of residence (**Supplementary Table 6**). When we explored differences in survival across hospitals these appeared to be more dramatic. The highest performing hospital in the country had a 5-year crude survival of 77.7% (95%CI 58.8, 88.7), while the corresponding estimate for the lowest performing hospital was 24.1% (95%CI 10.7, 40.5; **Supplementary Table 7**). We also found that for seven hospitals 10% or more of deaths occurred in the induction phase of treatment (induction mortality).

Demographic and clinical characteristics were similar between patients treated in their home state and those who migrated for treatment. However, a higher proportion of children who migrated for treatment were high-risk (72.5% vs. 68.9% treated in the home state; **Supplementary Table 8**). Nevertheless, after adjustment for several factors, survival was higher among children who migrated for treatment as compared to those who remained in their state of residence (HR=0.89; 95%CI 0.81, 0.98; **Table 2**). We found mortality to be higher in patients who were not treated in a paediatric hospital or in a facility without a paediatric oncology/haematology specialist. Multivariable-adjusted HRs were 1.18 (95% CI: 1.09, 1.26) for treatment in a non-paediatric specialty hospital and 2.17 (95%CI: 1.62, 2.90) for treatment by a non-paediatric specialist. Patient volume of the treating hospital was associated to ALL survival. Patients treated in hospitals that treated <20 patients in the previous year had a 22% higher mortality relative to those treated in a high patient-volume hospital (≥36 patients; HR= 1.22 (95%CI: 1.13, 1.32).

**Table 2.**
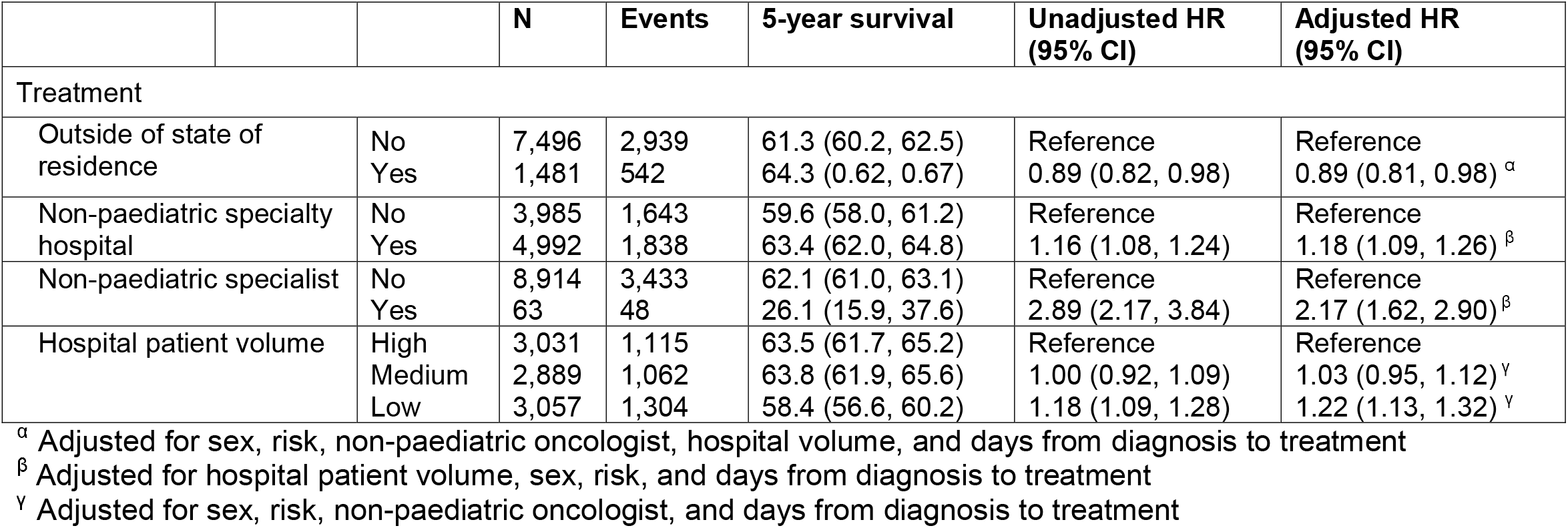
Hazard ratios and 95% confidence intervals (95% CI) for mortality comparing patient groups of childhood ALL treated under *Seguro Popular*’s policy for financing high-cost treatments, 2005-2015.

## Discussion

In this 10-year analysis of the health outcomes of a cancer care financing policy that was part of Mexico’s 2003 health reform, we found that access to treatment for childhood ALL doubled and by 2015 the policy financed the vast majority of estimated ALL cases for children without social security. We observed marked heterogeneity in ALL survival across states and hospitals, and identified factors amenable to intervention that could reduce these disparities.

Between 2005 and 2015 Mexico expanded funding, coverage, and access to childhood ALL treatment protocols, but survival did not improve over time in children without social security. An estimated 1,923 new cases of childhood ALL occur in Mexico every year.^15^ Based on 57.2%^2^ population coverage by *Seguro Popular*, we estimated that 1,100 of yearly cases happen in children without social security. Thus, our results suggest that by 2015, *Seguro Popular* financed the treatment of more than 95% of previously uninsured children with ALL (1,070 out of 1,100 estimated cases). As financing for childhood ALL expanded in the study period, more facilities were accredited for treatment and more children were treated in their home state, potentially reducing indirect costs of treatment (e.g., travel) and the risk of treatment abandonment.

Although some progress may have been achieved, based on survival for ALL, Mexico underperformed in survival for childhood ALL when compared to other countries in Latin America with similar income levels and healthcare systems. The 5-year survival (61.8%) was 10 percentage points below Chile and Argentina^5 6^—a stark indicator of underperformance. And contrary to expectations that sustained financing would result in improvements in quality of care, our analysis did not provide evidence for an increasing trend in survival over time. While overall the proportion of high-risk patients is similar to a previous study in Mexico City,^16^ this observation could be explained by an increase in the proportion of high-risk patients over time. The lack of improvement in survival is consistent with CONCORD-3 ALL survival estimates for Mexico for the same period.^3^ In contrast, that study found that Argentina had significant improvements in survival while Chile did not. Nevertheless, relative to the period prior to its implementation or to an estimate for a major social security provider (placing ALL childhood survival closer to 50%), *Seguro Popular* may have achieved some progress in ALL survival.^17 18^

We observed wide health disparities evidence for across states and hospitals. Childhood ALL survival differed by more than 30 percentage points across Mexican states and by close to 50 percentage points when comparing different facilities. This heterogeneity could be partly attributable to patient selection (those seeking care outside of their state of residence may be different than those remaining in their home state) as well as differences in barriers for access and continuity of care including language, cultural perceptions, and distance to hospitals.^19 20^ However, this heterogeneity remained after standardizing survival by the national distribution of ALL risk categories and after limiting our analysis to children treated in their home states. Also, hospitals that treated ALL patients under *Seguro Popular* reported important challenges in infrastructure, workforce, and availability of antibiotics and oncologic drugs.^21^ We found a time to treatment (an important determinant of therapeutic success) that was almost two times longer than what is expected in high income countries.^22^ Our study underscores the importance of timely quality of care and provides further evidence that children and adolescents with ALL have better outcomes when treated by paediatric oncology-haematology specialists,^23^ in a paediatric speciality hospital, and in facilities with a high volume of patients.^24^

Mexico’s experience financing childhood ALL treatment under *Seguro Popular* provides lessons that add to previously proposed recommendations for global paediatric oncology and calls to strengthen healthcare delivery systems.^1 25^ First, financing is necessary to increase access to care. *Seguro Popular*’s FPGC provided funds for treatment of most children with ALL without social security, and thereby expanded access to treatment that would have otherwise been unavailable or would have resulted in catastrophic expenditures. However, FPGC funded expensive cancer drugs only and institutions (and families) may have been financially strained to provide complementary medical supplies and services, such as antibiotics, blood banking, and intensive care. A policy similar to Chile’s, which explicitly guarantees optimal access to diagnosis, management of treatment-related complications, and, when necessary, palliative care, would provide greater financial certainty to treating facilitites.^26^

Second, financing is not sufficient to improve health outcomes. The substantial disparities in survival across states and hospitals in Mexico and the observed relevance of institutional expertise, infrastructure, and specialized paediatric oncology/haematology care for survival in our study underscores the importance of focusing on quality of cancer care. Similarly in Colombia, despite universal coverage for all children with cancer, disparities between private and public insurance were in part attributable to limited infrastructure.^27^ However, while blood banks and intensive care units and access to treatment for infectious complications are fundamental for quality of care, continuous training of clinical personnel and capacity building are key for improving performance. One strategy that has been successful is twinning smaller facilities with high-quality paediatric care centres. The top two hospitals (based on 5-year survival) in our analysis are engaged in long-term twinning partnerships with United States institutions.^28 29^ In countries like Mexico, where some paediatric care facilities provide quality of care comparable to high-income countries, similar within-country twinning partnerships could be established.

Finally, collecting and analysing data and implementing easy-to-measure actionable key performance indicators are necessary to improve survival. *Seguro Popular* collected data on childhood ALL cases, but the information was not used to monitor program effectiveness. In addition to 5-year survival, we assessed early mortality (≤ 30 days after treatment initiation), a quality performance indicator of treatment-related complications.^30^ While the national average (5.3%) was much lower than Brazil and Egypt (∼20%),^31 32^ only 7 states were below the proposed international benchmark of 4%.^14^ This relatively easy-to-calculate indicator could have been used in quality improvement interventions by the Mexican Ministry of Health.

In January 2020 the Mexican government ended *Seguro Popular* and began a full review of related policies, including the payment mechanism for high-cost conditions. The government has an obligation to demonstrate that any policy changes do not undermine the expanded access to treatment achieved for childhood ALL under *Seguro Popular*. Based on the current analysis, the experience under *Seguro Popular* provides evidence for two important recommendations that will be useful to policy makers as Mexico reforms its national health system. First, accreditation of facilities that provide cancer care to children should be based on performance and not exclusively on infrastructure and availability of human resources (inputs). Establishment of national benchmarks and close monitoring of system-wide quality of care indicators, such as survival, could be used to identify underperforming facilities that require improvement through capacity building programs. Second, a facility-based information system on treatment provided and ALL outcomes is critical to improve performance. The current administration could continue effective administrative data collection on services for children with ALL. Otherwise, funding specifically-designated personnel to collect information in hospital-based registries would be necessary.

One important strength of our study is the use of a nationwide comprehensive reimbursement dataset for 12 of the 14 years that *Seguro Popular* financed high-cost treatment interventions. We evaluated the completeness and quality of this administrative dataset with external data and demonstrated the validity of linkage to mortality data. However, error is inevitable when relying on administrative data that were not designed for research or monitoring purposes. Our survival estimate is roughly 10 percentage points higher than those from a hospital-based registry^3^ and a retrospective medical record review.^7^ Underreporting of cases likely occurred in the hospital-based registry due to lack of human resources for registration or sufficient institutional incentive for reporting cases. Also, previous assessments may have included an important proportion of children who had been previously treated (i.e., in disease relapse) and at a higher risk for mortality. In our study, underreporting was unlikely because we used patient data submitted in order to process reimbursement claims by the treating hospitals and we explicitly excluded patients treated for relapse. Thus, our analysis probably provides more comprehensive inclusion of cases and more accurate survival estimates.

This study has limitations. First, relative to studies based on retrospective medical record reviews or prospective data collection, our analysis did not include clinical information for a heterogeneous condition that would be useful to characterize with more detail this group of patients (i.e., molecular studies) and evaluate event-free survival (e.g., relapse). Second, we were unable to adjust for indicators of socioeconomic status, which could explain some of the differences observed in our comparison of patient populations. However, variability in socioeconomic status may be somewhat restricted in the population covered by *Seguro Popular*, since most affiliates were not employees in the formal sector or government. Nevertheless, like all observational studies, confounding by unmeasured variables remains a possibility. Third, using childhood ALL survival to appraise *Seguro Popular*’s performance does not provide a complete assessment of the effectiveness of a comprehensive public insurance policy. However, childhood ALL survival may be an appropriate tracer of health system performance.^33^ The disease is well-defined and easy to diagnose, the absence of treatment results in death, and childhood ALL is highly sensitive to quantity and quality of care.^34^ Finally, our assessment of *Seguro Popular* is limited by the lack of robust information on ALL survival in the population without social security prior to implementation in 2003.

As countries move towards universal health care, using administrative information on treatment for specific cancers may be a practical and low-cost approach to monitor progress, assess performance, and evaluate universal health coverage programs like *Seguro Popular*. Research efforts directed at understanding the underlying social determinants, health system, and behavioral factors that explain disparities in survival could provide a robust basis for designing interventions to improve health system performance.

## Data Availability

Data is currently not available.

## Author contributions

Muñoz-Aguirre P and Huerta-Gutierrez R conducted data management and analysis, participated in the study design, participated in writing the first draft and edited and reviewed the manuscript. Zamora S supervised data management and analysis and reviewed and edited the manuscript. Mohar A, Zapata-Tarres M, Bautista-Arredondo S, Rivera-Luna R, and Reich MR participated in the study design, interpreted results, and reviewed and edited the manuscript. Vega-Vega L, Hernández-Ávila J, Morales-Carmona E, and Perez-Cuevas R supervised data management and analysis of validation data and data linkage and reviewed and edited the manuscript. Lajous M designed the study, supervised the analysis, wrote the first draft, reviewed and edited subsequent versions and is responsible for all the content. Lajous M is the manuscript’s guarantor.

## Transparency declaration

M. Lajous is the manuscript’s guarantor and affirms that this manuscript is an honest, accurate, and transparent account of the study being reported; that no important aspects of the study have been omitted; and that any discrepancies from the study as planned (and, if relevant, registered) have been explained.

## Dissemination declaration

We plan to disseminate the results to patient organisations.

## No Patient and Public Involvement

This research was done without patient involvement. Patients were not invited to comment on the study design and were not consulted to develop patient relevant outcomes or interpret the results. Patients were not invited to contribute to the writing or editing of this document for readability or accuracy.

